# The association of ethnicity and oncologic outcomes for oral cavity squamous cell carcinoma (OSCC)

**DOI:** 10.1101/2024.02.25.24303341

**Authors:** Kiana Mahboubi, Steven C. Nakoneshny, Khara Sauro, Samuel Roberts, Rob Hart, T Wayne Matthews, Joseph Dort, Shamir P Chandarana

## Abstract

**Objective:** To compare oncologic outcomes of South Asian (SA) patients treated at a regional cancer centre in Canada, for oral squamous cell carcinoma (OSCC), to the general population.

**Methods:** Adult patients who underwent primary surgical resection of OSCC +/- adjuvant treatment between 2009 and 2022 (N=697) were included. SA patients were identified using a validated method and compared to non-SA patients. Baseline characteristics, including betel nut consumption, were compared, and disease-specific survival (DSS) and recurrence-free survival (RFS) were evaluated using Kaplan-Meier methods, with median follow-up time of 36.4 months [SD 31.02]. Cox proportional hazard regression models adjusted for potential confounders. A p-value < 0.05 was considered statistically significant.

**Results:** SA patients (9% of cohort, n = 64) were significantly younger and had lower rates of smoking and alcohol consumption compared to non-SA patients. There were no differences in tumor characteristics or the use of adjuvant radiation. SA patients had a two-fold higher risk of recurrence and significantly worse disease-specific survival, even after adjusting for stage and high-risk features [RFS: HR 2.01(1.28 - 3.14), DSS: HR 1.79(1.12 - 2.88)]. The consumption of betel nut was not associated with outcomes.

**Conclusion:** To our knowledge, this study is the first to compare the oncological outcomes of SA patients with OSCC to non-SA patients. SA patients had significantly worse outcomes, even after controlling for known predictors of recurrence and disease-specific survival. These findings can inform personalized treatment decisions and influence public health policies when managing patients with different ethnic backgrounds.

## Introduction

Head and neck cancer is the sixth most common cancer worldwide and is the most common neoplasm in South Asia (1). Oral cavity cancers account for 30% of all head and neck cancers and 90% of these are oral squamous cell carcinomas (OSCCs) (2,3). These cancers include tumours of the floor of the mouth, anterior tongue, alveolar ridge, retromolar trigone, the hard palate, and the buccal mucosa. According to the World Health Organization, of the 267,000 newly diagnosed OSCCs worldwide, close to 40% occurred in South Asia (India, Pakistan, Bangladesh, and Sri Lanka). Furthermore, oral cancer incidence and mortality rates in South Asia are almost twice those of global rates (4).

There are a variety of factors that are known to contribute to the development of OSCCs including the consumption of tobacco and alcohol, dental trauma, and human papilloma virus (5). The predominance of OSCCs in South Asia is often attributed to the use of betel nut (6), which can be consumed in a variety of ways but is most often dried and ground into a powder and wrapped in a package known as betel quid or pan, comprised of a mixture of slaked lime, flavouring, and tobacco. Its use is influenced by social acceptability, religious beliefs, and stimulant properties. Betel quid is often stored inside the cheek for hours similar to chewing tobacco. The slaked lime, most often used in India, is particularly problematic as it causes oxidative DNA damage and local mucosal abrasion creating deeper exposure to the carcinogenic components (7).

Although there have been improvements in the quality of life of patients with OSCC, both the disease itself as well as its treatment remain morbid with a 5-year overall survival rate of 50 to 60% (8). Current treatments include surgical resection, radiotherapy, chemotherapy, or a combination of these modalities. The primary treatment for early-stage (stage I and II) and advanced-stage (stage III and IV) OSCC is surgical resection of the tumor (9). Previous North American studies have looked at the differences in head and neck cancer disease outcomes according to ethnicity, showing large disparity in mortality rates among African American versus white patients in the United States (10-12). These differences have been attributed to a combination of tumour stage at time of presentation, access to healthcare, and exposure to carcinogens. Nichols *et al*., reported that even after controlling for tumour stage at time of presentation, African American patients had poorer outcomes, suggesting other intrinsic and extrinsic factors such as genetics and socioeconomic status influence survival in OSCC (13). Arbes *et al*., accounted for socioeconomic status, which resulted in elimination of the survival disadvantage observed among black patients (14). To date, however, there has been little research that compares oncological outcomes of patients of South Asian (SA) ethnicity with OSCCs compared to other ethnicities.

Our regional cancer centre, based in Calgary, Alberta, Canada, treats a high volume of OSCC, in patients with diverse ethnic backgrounds. SA patients represent a significant proportion of patients treated. Given the increased incidence and mortality rates of patients with OSCC in India, the objective of this study was to characterize oncological outcomes among patients of SA ethnicity within our centre and compare these outcomes to patients with OSCC who are not of SA ethnicity.

## Methods and Material

### Patient Selection

Adult patients (age 18 or older) that underwent curative intent primary surgical resection of OSCC at a regional cancer centre were included in the cohort. The set up of this study is that of a regional cancer centre where all head and neck cancers are managed by a multidisciplinary team with expert training and extensive experience. Patient data (n=697) was prospectively collected, and included all patients treated between 2009 to 2022. Patients with recurrent OSCC, a second primary malignancy, a synchronous primary malignancy, or who did not receive surgery as a primary treatment modality were excluded (Fig 1). Variables collected included: patient demographics, risk factors (ethnicity, age, gender, smoking and alcohol status, betel nut use), pathologic data (AJCC 8^th^ edition TNM staging (15) including presence of extranodal extension [ENE], lymphovascular invasion [LVI], perineural invasion [PNI]), and treatment (adjuvant radiation and/or chemotherapy).

Patients of SA descent were identified using a multistep approach. First, a previously constructed and validated SA surname list (16) was used to assign SA ethnicity to the patient population. The validated SA surname list was then linked with the patient database to generate a SA patient list. In addition, to ensure the accuracy of the cohort of SA patients, the final study cohort was manually entered into a surname’s origin website (17). This list was reviewed by two researchers with SA/middle eastern backgrounds to generate a final list of SA patients. This method has been used successfully to identify ethnic groups in other studies (18, 19).

### Statistical Analysis

We interrogated a prospectively collected database of all patients treated for OSCC at the Calgary regional cancer centre. Patient factors, tumor factors, treatment factors and outcomes were analyzed and compared between the SA and non-SA groups.

Categorical outcomes were compared between groups using chi-square and continuous outcomes were compared using Student’s t-test. Primary outcomes were recurrence-free survival (RFS) and disease specific survival (DSS). RFS was defined as time from surgery to time of last follow-up or local/regional/distant recurrence, whichever came first. DSS was defined as time from surgery to time of last follow-up or death related to OSCC. Difference in RFS and DSS between patients of SA ethnicity and those of non-SA ethnicity were determined by comparing the time-to-event (Kaplan-Meier survival curves) using a log-rank test statistic. These were censored at 3 years of follow-up. Time-to-event outcomes were then adjusted for variables that potentially modified or confounded the relationship between the outcome and exposure (social habits, stage, high risk features on surgical pathology, type of adjuvant treatment) using Cox proportional hazards (PH) regression models. These variables were, in part, based on clinical relevance, having been shown in the literature to potentially modify the oncologic outcomes (20-24). Variables that had a p-value of <0.20 in the univariate model were included in the multivariable model. Stepwise selection methods were used to develop the final models. A p-value < 0.05 was considered statistically significant. Owing to the prospective and rigorous nature of the data collection, missing data was infrequently seen; when missing data was encountered, it was excluded from analysis. Betel nut consumption was not prospectively collected, but due to the published association between consumption and carcinogenesis, a chart review was performed, and where possible, betel nut consumption was collected. A sub-analysis of only patients of SA ethnicity was performed to look for association between betel nut consumption and oncologic outcome (DSS and RFS), by comparing the time-to-event (Kaplan-Meier survival curves) of those that did and did not consume betel nut, using a log-rank test statistic.

Statistical Analysis was performed using Stata, version 14 (25). The study was approved by the University of Calgary Conjoint Health Research Ethics Board.

## Results

Using the previously described approach for selecting SA patients, we identified 64 SA patients and 632 non-SA patients, which served as the comparison group. We were unable to classify 1 patient as either SA or non-SA (Figure 1). Of the 697 patients that were included, 9% (n=64) were of SA ethnicity. Table 1 describes the cohort by patient and tumour characteristics stratified by ethnicity. The median follow-up time was 36.4 months (SD=31.02). SA patients were significantly younger and were less likely to smoke or drink. There were no differences in tumor pathologic characteristics (T-stage, N-stage, ENE, LVI, PNI), nor in the use of adjuvant radiation between the SA and non-SA patients.

**Table 1.**
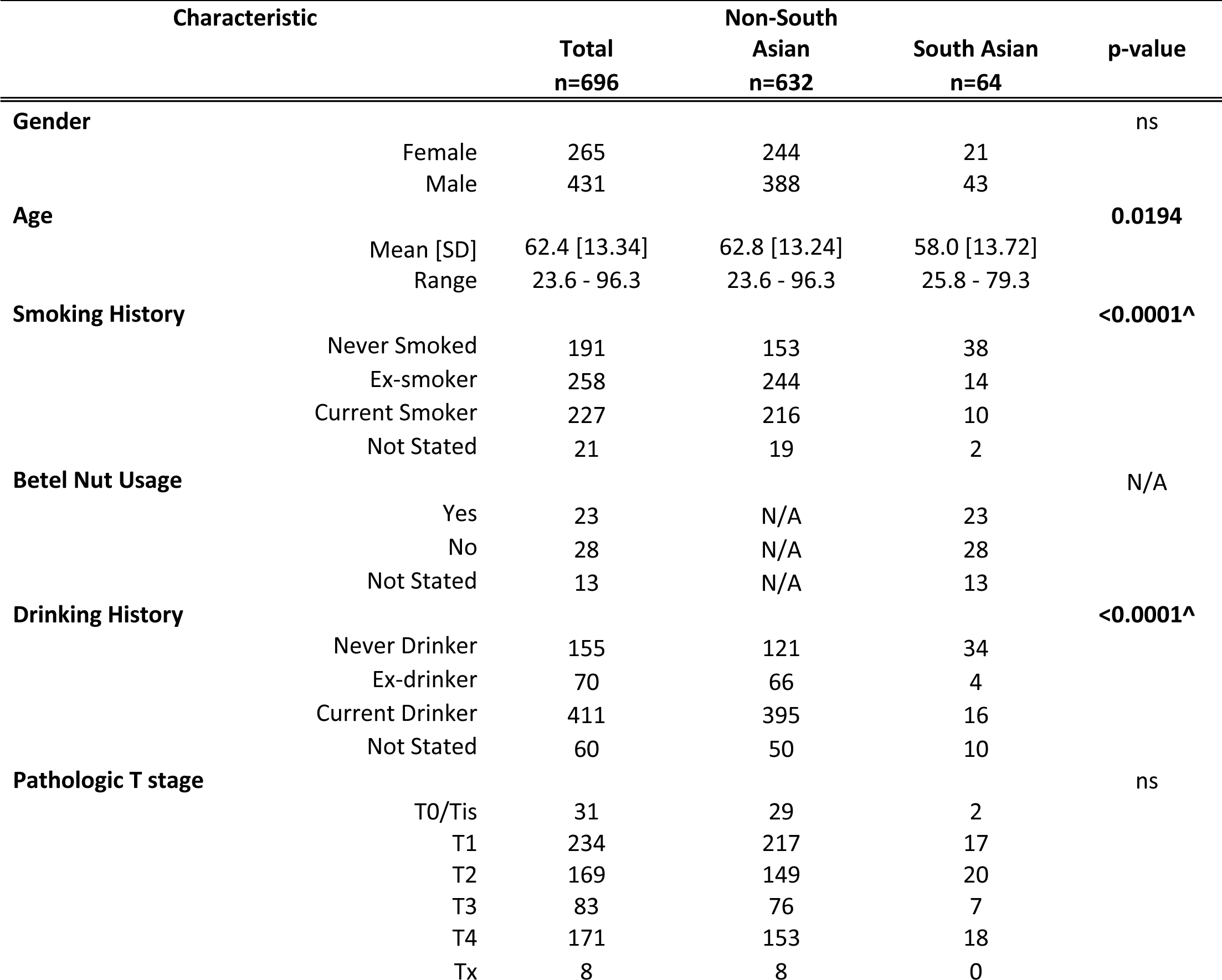
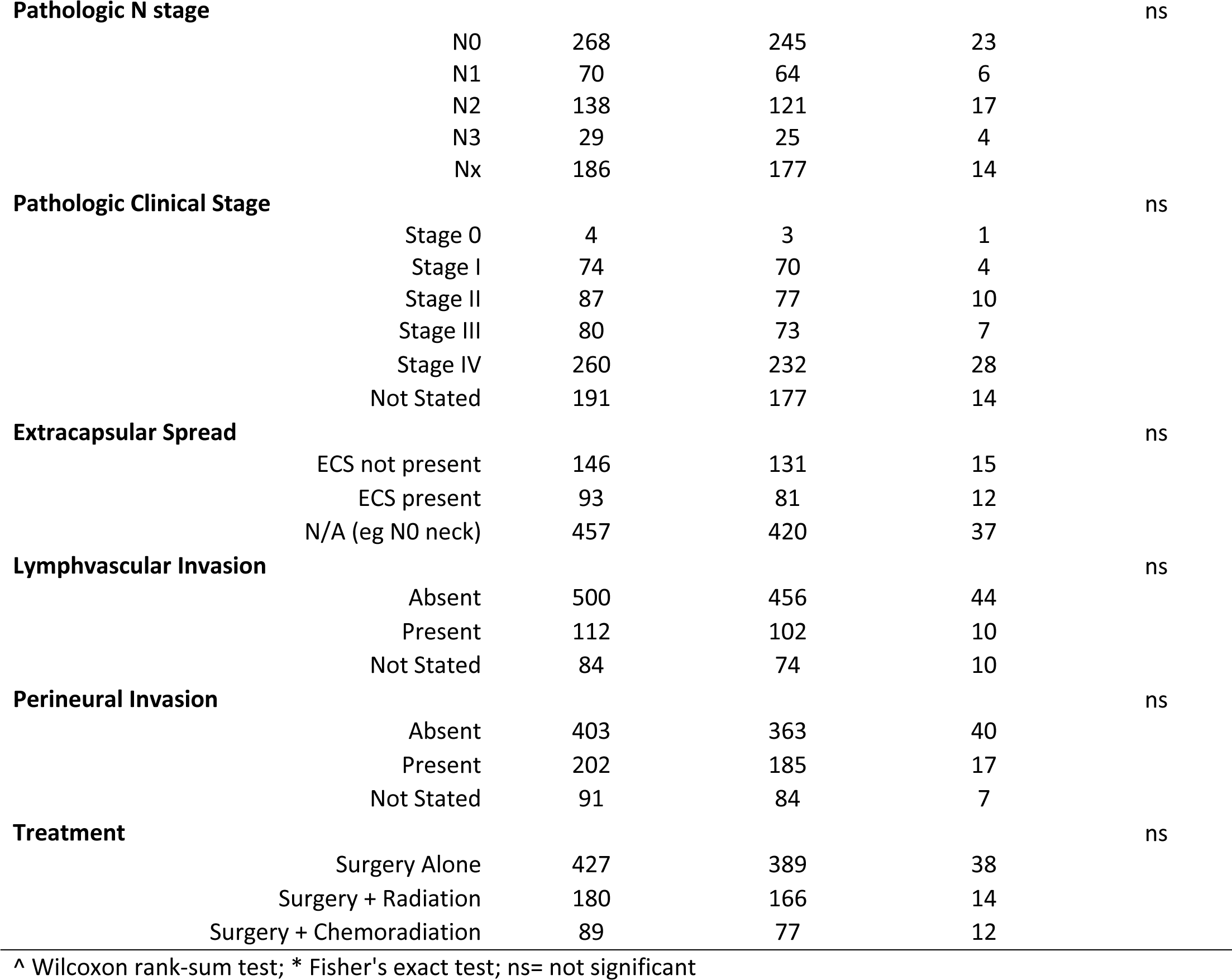
Patient Demographics.

| Characteristic |  | Total<br>n=696 | Non-South<br>Asian<br>n=632 | South Asian<br>n=64 | p-value |
| --- | --- | --- | --- | --- | --- |
| **Gender** |  |  |  |  | ns |
|  | Female | 265 | 244 | 21 |  |
|  | Male | 431 | 388 | 43 |  |
| **Age** |  |  |  |  | **0.0194** |
|  | Mean [SD] | 62.4 [13.34] | 62.8 [13.24] | 58.0 [13.72] |  |
|  | Range | 23.6 - 96.3 | 23.6 - 96.3 | 25.8 - 79.3 |  |
| **Smoking History** |  |  |  |  | **<0.0001^** |
|  | Never Smoked | 191 | 153 | 38 |  |
|  | Ex-smoker | 258 | 244 | 14 |  |
|  | Current Smoker | 227 | 216 | 10 |  |
|  | Not Stated | 21 | 19 | 2 |  |
| **Betel Nut Usage** |  |  |  |  | N/A |
|  | Yes | 23 | N/A | 23 |  |
|  | No | 28 | N/A | 28 |  |
|  | Not Stated | 13 | N/A | 13 |  |
| **Drinking History** |  |  |  |  | **<0.0001^** |
|  | Never Drinker | 155 | 121 | 34 |  |
|  | Ex-drinker | 70 | 66 | 4 |  |
|  | Current Drinker | 411 | 395 | 16 |  |
|  | Not Stated | 60 | 50 | 10 |  |
| **Pathologic T stage** |  |  |  |  | ns |
|  | T0/Tis | 31 | 29 | 2 |  |
|  | T1 | 234 | 217 | 17 |  |
|  | T2 | 169 | 149 | 20 |  |
|  | T3 | 83 | 76 | 7 |  |
|  | T4 | 171 | 153 | 18 |  |
|  | Tx | 8 | 8 | 0 |  |
| **Pathologic N stage** |  |  |  |  | ns |
|  | N0 | 268 | 245 | 23 |  |
|  | N1 | 70 | 64 | 6 |  |
|  | N2 | 138 | 121 | 17 |  |
|  | N3 | 29 | 25 | 4 |  |
|  | Nx | 186 | 177 | 14 |  |
| **Pathologic Clinical Stage** |  |  |  |  | ns |
|  | Stage 0 | 4 | 3 | 1 |  |
|  | Stage I | 74 | 70 | 4 |  |
|  | Stage II | 87 | 77 | 10 |  |
|  | Stage III | 80 | 73 | 7 |  |
|  | Stage IV | 260 | 232 | 28 |  |
|  | Not Stated | 191 | 177 | 14 |  |
| **Extracapsular Spread** |  |  |  |  | ns |
|  | ECS not present | 146 | 131 | 15 |  |
|  | ECS present | 93 | 81 | 12 |  |
|  | N/A (eg N0 neck) | 457 | 420 | 37 |  |
| **Lymphovascular Invasion** |  |  |  |  | ns |
|  | Absent | 500 | 456 | 44 |  |
|  | Present | 112 | 102 | 10 |  |
|  | Not Stated | 84 | 74 | 10 |  |
| **Perineural Invasion** |  |  |  |  | ns |
|  | Absent | 403 | 363 | 40 |  |
|  | Present | 202 | 185 | 17 |  |
|  | Not Stated | 91 | 84 | 7 |  |
| **Treatment** |  |  |  |  | ns |
|  | Surgery Alone | 427 | 389 | 38 |  |
|  | Surgery + Radiation | 180 | 166 | 14 |  |
|  | Surgery + Chemoradiation | 89 | 77 | 12 |  |

^ Wilcoxon rank-sum test; \* Fisher's exact test; ns= not significant

### Recurrence free survival

Univariate Kaplan-Meier analysis revealed that patients of SA ethnicity had worse RFS [HR=2.35 (1.51–3.65)] than those of non-SA ethnicity (Figure 2A). RFS for SA patients at 3-year follow up was 52%, compared to 76% in the non-SA group (p<0.01). Pathologic characteristics were significantly associated with worsened RFS, regardless of ethnicity: advanced T-stage [HR=2.17 (1.55–3.04)], node positivity [HR= 2.98 (1.99–4.45)], presence of ENE [HR=5.39 (3.57– 8.14)], advanced clinical stage [HR = 5.14 (2.82–9.36)], presence of LVI [HR = 2.93 (2.00–4.29)], and presence of PNI [HR = 1.78 (1.24–2.55)] (Figure 2A).

After adjusting for the above-mentioned covariates on multivariable analysis, SA ethnicity [HR=2.01 (1.28 - 3.14)], advanced T-stage [HR =1.46 (1.02 - 2.10)], nodal positivity [HR=2.65 (1.77 - 3.99)], and presence of ENE [HR=4.32 (2.75 - 6.78)] were all associated with worsened RFS (Table 2).

**Table 2:** Cox Regression Analysis for Recurrence Free Survival.

| Variable | Univariate Analysis |  | Multivariable Analysis |  |
| --- | --- | --- | --- | --- |
|  | Hazard ratio<br>[95% CI] | p-value | Hazard ratio<br>[95% CI] | p-value |
| Ethnicity, non-SA vs SA | 2.35 [1.51 - 3.65] | **<0.0001** | 2.01 [1.28 - 3.14] | **0.002** |
| Pathologic T Classification, pT0-pT2 vs pT3-pT4 | 2.17 [1.55 - 3.04] | **<0.0001** | 1.46 [1.02 - 2.10] | **0.04** |
| Pathologic Node Positivity, pN0 vs<br>pN+ | 4.81 [3.08 - 7.51] | **<0.0001** | — | — |
| Clinical Stage, I/II vs III/IV | 5.14 [2.82 - 9.36] | **<0.0001** | — | — |
| Lymphovascular Invasion, no vs<br>yes | 2.93 [2.00 - 4.29] | **<0.0001** | — | — |
| Perineural Invasion, no vs yes | 1.78 [1.24 - 2.55] | **0.002** | — | — |
| Extracapsular Spread (ref: N0) |  |  |  |  |
| N+, ECS— | 2.98 [1.99 - 4.45] | **<0.0001** | 2.65 [1.77 - 3.99] | **<0.0001** |
| N+, ECS+ | 5.39 [3.57 - 8.14] | **<0.0001** | 4.32 [2.75 - 6.78] | **<0.0001** |
| Treatment Modality (ref: Surgery<br>Alone) |  |  |  |  |
| Surgery + Radiation | 2.30 [1.57 - 3.36] | **<0.0001** | — | — |
| Surgery + Chemoradiation | 2.84 [1.83 - 4.34] | **<0.0001** | — | — |

### Disease-specific survival

On univariate Kaplan-Meier analysis, SA ethnicity was associated with worsened disease-specific survival [HR=2.14 (1.34–3.42)]. DSS for SA patients at 3-year follow up was 59%, compared to 77% in the non-SA group (p<0.01). Other negative prognostic features included: advanced T-stage [HR=2.37 (1.66–3.36)], nodal positivity [HR=3.70 (2.38–5.74)], presence of ENE [HR=8.04 (5.24– 12.35], advanced clinical stage [HR = 7.67 (3.73–15.78)], presence of LVI [HR = 3.11 (2.11–4.57)], and presence of PNI [HR = 2.20 (1.53–3.18)] (Figure 2B).

After adjusting for the above-mentioned covariates on multivariable analysis, SA ethnicity [HR=1.79 (1.12–2.88)], advanced T-stage [HR = 1.64 (1.10–2.43)], node positivity [HR=3.97 (2.44–6.45)], and presence of ENE [HR=16.40 (9.00–29.91)] predicted worsened DSS (Table 3).

**Table 3:** Cox Regression Analysis for Disease Specific Survival.

| Variable | Univariate Analysis |  | Multivariable Analysis |  |
| --- | --- | --- | --- | --- |
|  | Hazard ratio<br>[95% CI] | p-value | Hazard ratio<br>[95% CI] | p-value |
| Ethnicity, non-SA vs SA | 2.14 [1.34 - 3.42] | **0.001** | 1.79 [1.12 - 2.88] | **0.016** |
| Pathologic T Classification, pT0-pT2 vs pT3-pT4 | 2.37 [1.66 - 3.36] | **<0.0001** | 1.64 [1.10 - 2.43] | **0.015** |
| Pathologic Node Positivity, pN0<br>vs pN+ | 6.10 [3.75 - 9.92] | **<0.0001** | — | — |
| Clinical Stage, I/II vs III/IV | 7.67 [3.73 -<br>15.78] | **<0.0001** | — | — |
| Lymphovascular Invasion, no vs<br>yes | 3.11 [2.11 - 4.57] | **<0.0001** | — | — |
| Perineural Invasion, no vs yes | 2.20 [1.53 - 3.18] | **<0.0001** | — | — |
| Bone Invasion, no vs yes | 1.80 [1.11 - 2.92] | **0.017** | — | — |
| Extracapsular Spread (ref: N0) |  |  |  |  |
| N+, ECS— | 3.70 [2.38 - 5.74] | **<0.0001** | 3.97 [2.44 - 6.45] | **<0.0001** |
| N+, ECS+ | 8.04 [5.24 -<br>12.35] | **<0.0001** | 16.40 [9.00 -<br>29.91] | **<0.0001** |
| Treatment Modality (ref: Surgery<br>Alone) |  |  |  |  |
| Surgery + Radiation | 2.28 [1.53 - 3.41] | **<0.0001** | 0.85 [0.53 - 1.36] | ns |
| Surgery + Chemoradiation | 3.14 [2.00 - 4.94] | **<0.0001** | 0.27 [0.14 - 0.53] | **<0.0001** |

ns= not significant

### Pattern of failure in those that recurred

Among the 193 patients that developed a recurrence, 86% of patients of SA ethnicity recurred either locally or regionally, compared to the 87% in the patients of non-SA ethnicity. Conversely, 14% of patients of SA ethnicity and 13% of patients of non-SA ethnicity had distant recurrence. The difference in pattern of failure was not significant.

### Betel nut use in SA patients

Chart review of the 64 SA patients revealed that 23 used betel nut, 28 did not use betel nut, and 13 did not have betel nut use reported. Univariate analysis of just the SA patients did not predict a difference in RFS or DSS between those who used betel nut and those who did not (Figure 3).

## Discussion

This study demonstrated that patients of SA ethnicity had significantly worse survival outcomes and were twice as likely to recur compared to non-SA patients even after accounting for other known factors contributing to poor oncological outcomes. The SA community is one of the largest and fastest growing minority groups in Canada based on the 2021 census (26). Considering the migration patterns within the South Asian community and their correlation with the prevalence of OSCC, it’s important to acknowledge the potential impact on health trends in Canada. Understanding these patterns, can help promote public health initiatives and ensure that the unique needs of the South Asian population are met, fostering a more inclusive and supportive environment. Studies conducted in Malaysia (27,28), UK (29), Australia (30), and South Africa (31) indicate that individuals of SA heritage are at higher risk of oral cancer than the non-SA population in those countries. A study in British Columbia, which has one of the highest South Asian immigrant populations in Canada, demonstrated a relative risk of developing OSCC of 1.33 and 1.66 for South Asian men and women, respectively, as compared with the non-SA population (32). Anecdotally, the senior authors at our centre, who treat high volumes of OSCC, felt that year over year, SA patients represented a disproportionately larger ethnicity group than other ethnicities, and were recurring more frequently. The decision to formally evaluate outcomes based on SA ethnicity was therefore born out of concern for a potentially at-risk population, in hopes to validate the need for a more tailored approach to treating this group of patients, as well as to build awareness and inform public health policy.

This study uniquely evaluates differences in oncological outcomes of patients of SA ethnicity affected by OSCCs compared to patients of non-SA ethnicity. This results suggest that, in Canada, SA ethnicity is an independent predictor of recurrence-free and worse disease-specific survival, when compared to non-SA patients. The study is conducted within the framework of a regional cancer center, where a dedicated multidisciplinary team, possessing expert training and extensive experience, oversees the management of all head and neck cancers. This setup intentionally minimizes bias for selection for more complicated cases. Stage of diagnosis is regarded as one of the most important predictors of oral cancer survival with a significantly improved 5-year survival rate for early-stage disease (71.4%) than for late-stage disease (21.8%) (33). In this study, when comparing baseline characteristics of SA and non-SA patients, oncological parameters such as T-Stage, N-stage, ENE, LVI and PNI were similar which suggests they were not the primary drivers of differences in recurrence and worse survival in SA patients. This association was also found in multivariable analysis. The similarity in these variables between groups also infers that there are no discrepancies in access to care, which would likely lead to a delayed presentation, with more advanced disease at time of presentation. Although, the initial presenting burden of disease is an imperfect tool to estimate access to care, in a universal health care system, issues such as treatment delay, is relatively uniform across all sub-populations. Interestingly, features that are typically protective against developing OSCC such as younger age and non-smoking status were more common in SA patients. Despite this, being SA resulted in a two-fold increase in recurrence and decrease in OSCC survival. As expected, patients with advanced T stage (T3/4), nodal disease and ENE had both worsened DSS and RFS, validating that the patient cohort in this study is representative of the greater head and neck cancer population. Disease stage at time of diagnosis has been previously identified as a causative factor in poorer oncological outcome in minority groups (34). In this study, even after controlling for disease stage, SA ethnicity remained as an independent factor in predicting survival and recurrence.

Several factors potentially contribute to the poorer outcomes of OSCC in SA patients. One of the major factors is thought to be the use of betel nut in the SA culture, which is commonly consumed in the form of betel quid. Although the added tobacco plays a significant role in the development of OSCCs, studies have suggested that betel products, which contain arecoline and 3-(methylnitrosamino) propionitrile, may have an independent carcinogenic effect (35) resulting in malignant transformation of oral submucosal fibrosis (OSMF). The potential for malignant transformation resulting in OSCC has been reported to be as high as 7 to 13% (36). Although the exact mechanism is not well understood, betel nut is thought to induce *c-jun* proto-oncogene expression in human mucosal fibroblasts (37). The fibrosis itself can result in decreased vascularity and hypoxia thus mediating mutated cell divisions (38). This study did not find any differences on either recurrence or survival among patients of SA ethnicity based on betel nut use; however, given that betel nut consumption was not consistently reported in patient charts, further investigation with larger numbers and more consistent reporting is required to verify this finding.

Pattern of recurrence describes whether a patient recurs locally, regionally, or at a distant site. Given that betel nut is often stored inside the cheek for prolonged periods of time, one would expect SA patients to recur locoregionally if betel nut use is the causative factor. In our study population, the majority of recurrences in the SA population were local/regional however, the proportion of patients that recurred locoregionally was not significantly different between SA and non-SA patients. This suggests that regardless of the causative agent for OSCC, recurrences are more likely to manifest as local/regional rather than distant disease.

A strength of this study relies on the rigorous, prospective collection of data including patient related factors, tumor factors, treatment factors and oncologic outcomes. A limitation is the relatively small sample size within the SA cohort. A larger multicentre study could yield more definitive results on this issue. Given the absence of race or ethnicity information in many databases used in health research, surnames are frequently employed as a proxy when investigating healthcare patterns within ethnic populations. Despite this, a limitation arises in potentially excluding individuals of South Asian background who may have undergone name changes. However, it should be noted that in Canada, according to the 2021 census, 76% of South Asians were recent immigrants born outside of Canada (26), and therefore, the likelihood of experiencing a name change is low. This study also did not differentiate between SA immigrants and Canadian born people of SA ethnicity (first and subsequent generations) which hinders the ability to distinguish between the role of environment versus inheritance on the findings. There are important factors related to immigration status that may mediate the findings of this study, some of which include betel nut use, environmental factors and genetic factors. Although, there is no specific data on the use of betel nut in Canada, UK and Australian data shows that it is commonly used among first generation immigrants and its use is reduced in subsequent generations (39). However, in younger generations, the form of consumption is tilting towards ingestion which may result in higher rates of esophageal cancers (39). As such, a more granular study on time since immigration and mode of betel nut use would be informative. In addition to environmental factors, genetics can also play a role in the poor outcomes observed in South Asian patients with OSCC. However, no specific genes have been identified as predisposing factors for OSCC in the SA population and the hereditary factors that contribute to the disease are largely unknown. While most cases of OSCC occur sporadically, certain families with a high preponderance of the disease have been found to carry oncogenes such as *VAV2* and *IQGAP1* with an autosomal dominance penetrance (40). Moreover, with the emergence of precision medicine, there has been a growing interest in identifying diagnostic and prognostic biomarkers for OSCC. One such biomarker is microRNA (miRNA), a large group of small single-stranded non-coding endogenous RNAs that play a role in post-transcriptional gene regulation. Upregulation of miRNAs is thought to contribute to OSCC resistance to chemoradiation and recurrence (41). Our centre is involved in banking the tumors of all consecutive OSCC patients and intends to explore the potential molecular basis of poor OSCC outcomes in the SA population.

The findings of this study reveal that patients with SA ethnicity experience significantly poorer outcomes compared to the non-SA cohort, even after accounting for other predictors of poor outcome. To gain a more comprehensive understanding of the intricate interplay between ethnicity, environmental exposure, and genetic predisposition in OSCC outcomes, further research is imperative. Such research could pave the way for personalized treatment decisions and patient counseling, as well as the formulation of public health policies that serve to assist vulnerable and at-risk populations.

## Conclusion

Patients of SA ethnicity had worse outcomes (higher risk of recurrence and worsened survival) than patients with non-SA ethnicity, even after controlling for other known predictors of poor outcome in OSCC. These finding can inform more individualized treatment decision making and impact public health policy when serving heterogeneous patient populations.

## Data Availability

All data produced in the present study are available upon reasonable request to the authors

